# Early Feasibility Study of Sensing-enabled Ventral Capsule Deep Brain Stimulation in 10 Participants with Intractable Obsessive-Compulsive Disorder

**DOI:** 10.64898/2026.04.24.26351318

**Authors:** Sameer A. Sheth, Nicole R. Provenza, Sarah Soubra, Thomas Hamre, Ben Shofty, Garrett P. Banks, Nisha Giridharan, Faiza Momin, Greg Vogt, Sarah McKay, Michelle Avendano Ortega, Luke Jumper, Andrew D. Wiese, Eric A. Storch, Wayne K. Goodman

## Abstract

**Objective:** To address the limitations of current trial-and-error programming strategies in deep brain stimulation (DBS) for refractory obsessive-compulsive disorder (OCD), we implanted patients with sensing-capable DBS devices to identify neural biomarkers that could provide objective feedback to the clinician about therapeutic efficacy.

**Methods:** We conducted an early feasibility study in 10 patients with severe, treatment-resistant OCD. All subjects received bilateral DBS leads targeting the ventral internal capsule (VC) and the second half of the cohort also received strip electrodes over the bilateral orbitofrontal cortex for recording only. All leads were connected to investigational, bidirectional DBS devices. After implantation, participants returned for scheduled programming visits to determine optimal stimulation parameters (Phase 1). In Phase 2, patients completed a course of exposure and response prevention (ERP) psychotherapy, and in Phase 3, patients underwent a double-blind discontinuation of DBS to test true vs. sham response. Phase 4 was an open-label follow-up. We administered standardized symptom scales throughout the study and used non-parametric repeated-measures analyses to analyze neuropsychological data.

**Results:** All patients elected to resume stimulation after a discontinuation phase. At the end of the study, the mean reduction in the Yale-Brown Obsessive-Compulsive Scale (Y-BOCS) was 22 points or 60% across all patients with 8 patients demonstrating full response (≥35% decrease in Y-BOCS). All participants also experienced reduced depression severity.

**Conclusions:** In patients with refractory OCD, we demonstrate excellent clinical response to VC DBS. We show feasibility of recording neural data both at home and in the clinic on board bidirectional DBS devices.

## Introduction

Obsessive-compulsive disorder (OCD) is a common, persistent, and oftentimes disabling disorder marked by unwanted and distressing thoughts (obsessions) and/or repetitive behaviors (compulsions)(1). OCD affects 2-3% of the US population(2) and is responsible for substantial functional impairment and increased risk of early mortality(3). First-line treatments for OCD are cognitive-behavioral therapy with exposure and response prevention (ERP)(4) and serotonin reuptake inhibitor (SRI) medications(5). Approximately 30-40% of patients fail to respond to either modality(6) few patients experience complete symptom resolution(7) and risk of relapse after treatment discontinuation remains significant(8) Non-invasive neuromodulation therapies like repetitive transcranial magnetic stimulation (rTMS) have shown modest benefit in non-treatment resistant OCD(9).

For the most severe and refractory cases, neurosurgical procedures can offer relief. In the past two decades, deep brain stimulation (DBS) targeting the ventral portion of the anterior limb of the internal capsule (ventral capsule, VC) has shown success in several trials(10–16) with a response rate of 66% according to a recent meta-analysis(17). In 2009, the FDA approved a Humanitarian Device Exemption (HDE) for DBS in intractable OCD based upon the device’s safety and probable benefit for up to 8,000 people a year in the US that would meet implantation criteria.

Despite growing evidence in support of DBS for refractory OCD, there is room for improvement in both clinical benefits and reduction of DBS-induced behavioral side effects, especially hypomania. A related limitation of DBS for OCD is that programming adjustments are largely based on acute beneficial effects on “mood” and “energy” as reported by patients and observed by clinicians. Immediate effects on core OCD symptoms are generally indiscernible during a programming session. Instead, parameters are adjusted in a trial-and-error fashion based on changes in OCD symptom severity. This temporal mismatch calls into question the causality of any particular adjustment and mystifies programming strategies. In contrast, adjustments of DBS parameters for Parkinson’s disease (PD) typically produce immediate changes in primary symptoms such as tremor or rigidity, thus facilitating programming and hastening clinical response.

To address these limitations, we conducted a study of DBS in OCD that employed a new generation of DBS devices with neural sensing capability(16). An objective measure that demonstrates target engagement, such as a neural biomarker, could provide this missing feedback and increase confidence in therapy delivery. It could also provide an objective measure of unwanted behavioral states such as DBS-induced hypomania(11,18). In appropriate circumstances such a biomarker could possibly be used for closed-loop applications as well, just as beta band power has been studied as a feedback signal for closed-loop DBS for PD(19).

The clinical backbone for this study, described herein, is an Early Feasibility Study in 10 patients with severe, treatment-resistant OCD. DBS was initially delivered open-label to identify patient-specific parameters. We introduced a course of ERP therapy after several months given the growing clinical(20) and mechanistic(18) evidence for synergy between DBS and ERP. To distinguish active from sham effects, patients then underwent blinded discontinuation of DBS therapy with specific rescue criteria.

## Methods

### Patient Selection

We used internationally accepted criteria(11,21) to determine candidacy for DBS, including diagnosis (OCD primary), chronicity (duration of >5 years), severity (Yale-Brown Obsessive-Compulsive Scale [Y-BOCS](22) >28), and refractoriness (to trials of at least 2 selective serotonin reuptake inhibitors, clomipramine, augmentation with an antipsychotic, and high-fidelity ERP).

### Surgery

All subjects received a pair of quadripolar DBS leads (Medtronic 3387, 1.5 mm gap between contacts) targeting the ventral portion of the internal capsule (VC). We used an implantation strategy we have previously described(13,16,18), creating trajectories to the VC both anterior to the anterior commissure (AC), such that the lead tip would border the ventral striatum (VC/VS), and posterior to the AC, such that the lead tip would border the bed nucleus of the stria terminalis (VC/BNST). We used intraoperative awake testing to adjudicate between trajectories, using stimulation-induced approach behaviors(13,18) (smiling, talkativeness, improved mood or energy) as a behavioral indicator of successful target engagement. Leads were connected via extensions to a subclavicular implanted pulse generator (IPG, Medtronic PC+S for P001-002, Medtronic RC+S for P003-010).

The second half of the cohort (P006-010) also received a pair of 4-contact strip electrodes (Medtronic 0913025) on the bilateral orbitofrontal cortex (OFC) to test the hypothesis that neural recordings from the OFC would provide additional predictive power of clinical state. Because of the stiffness of these electrodes, they could not be placed through the DBS burr hole and curved around the frontal cortex convexity but rather had to be placed along a straight-line trajectory via a burr hole at the floor of the anterior cranial fossa. Each DBS lead and OFC strip electrode was routed via extensions to an ipsilateral IPG (Medtronic RC+S) such that these 5 patients each received a pair of DBS leads, a pair of OFC strip electrodes, and two IPGs.

### Trial Design

All participants completed a consent/assessment visit between weeks -9 and -5 (relative to surgery). The immediate pre-surgical baseline visit (weeks -4 to -1) included a full neuropsychological battery, independent clinical evaluation, and neuroimaging. Two weeks after surgery, we performed a repeat neuropsychological exam and then initiated stimulation.

Following initial programming, subsequent programming visits occurred every 2 weeks for the next 2 months and then monthly. The first 6 months (Phase 1) were open-label programming to identify optimal stimulation parameters. We then added a 2-month, 15-session course of ERP(23) to test whether ERP following DBS could be synergistically effective (Phase 2, months 7-8). Phase 3 (month 9) consisted of double-blind discontinuation of DBS therapy to disambiguate true vs. sham response. Stimulation amplitude was tapered according to a fixed schedule over 4 weeks: 100% during week 1, 50% during week 2, and 0% stimulation for weeks 3 and 4. We decided to reinstate active DBS therapy at the end of month 9 if the patient had a relapse, defined as a 25% increase of the Y-BOCS over 2 consecutive visits compared to discontinuation baseline or a Clinical Global Impression-Improvement (CGI-I) score≥6. Phase 4 (months 10-18) was an open-label follow-up with a final neuropsychological battery at month 12.

We used non-parametric repeated-measures analyses to analyze the neuropsychological data due to small sample size and non-normal outcome distributions.

## Results

### Demographics

We enrolled 10 patients (5 female; 3 Hispanic/Latino) with severe OCD (Y-BOCS>28; Y-BOCS-II>36) (Table 1). Age of onset was in adolescence for most. The most common comorbidity was major depressive disorder (9/10). OCD symptoms spanned a range, with the most common being harm avoidance (7/10), contamination (7/10), and “not just right” feelings (6/10).

**Table 1.**
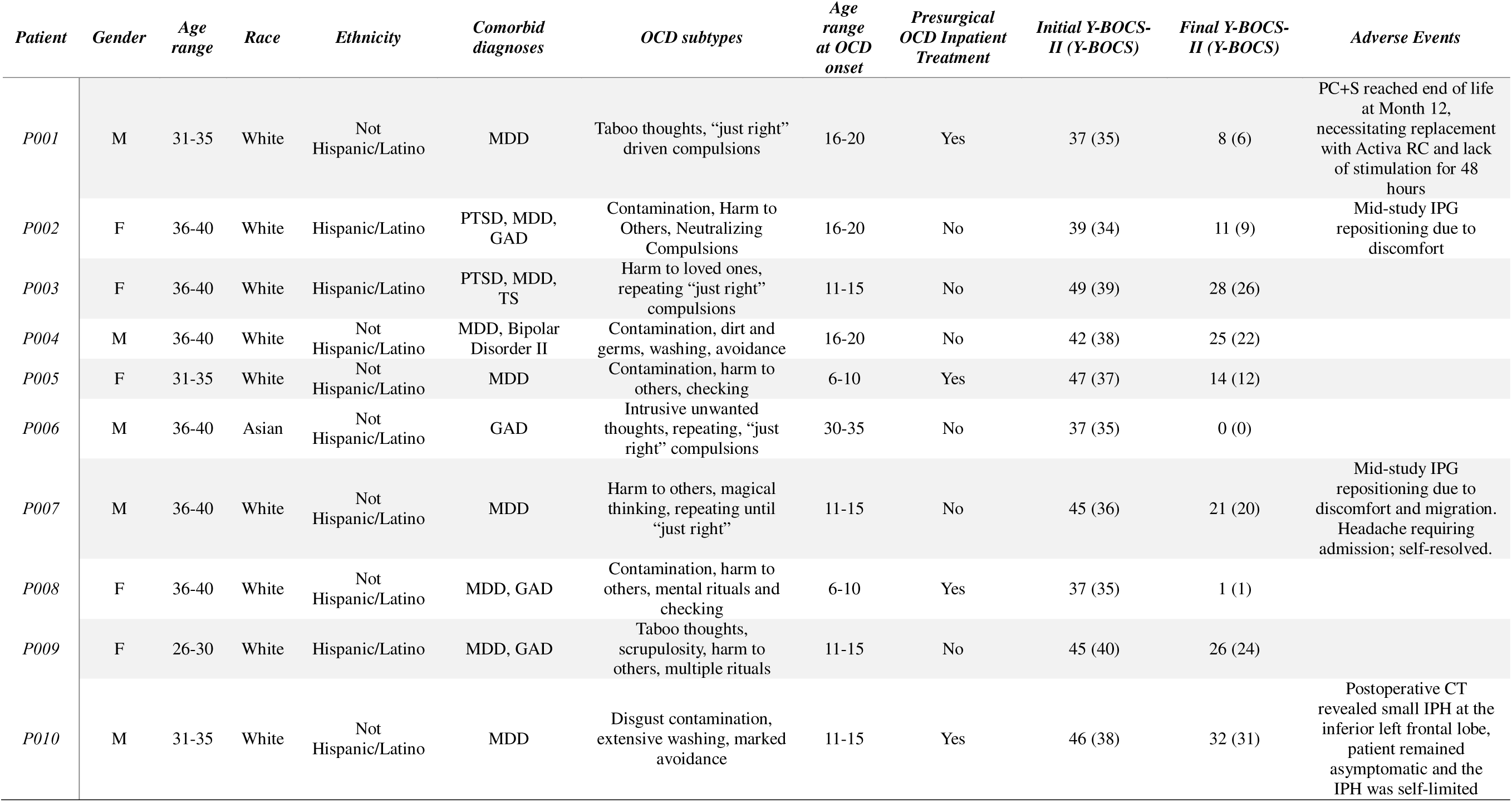
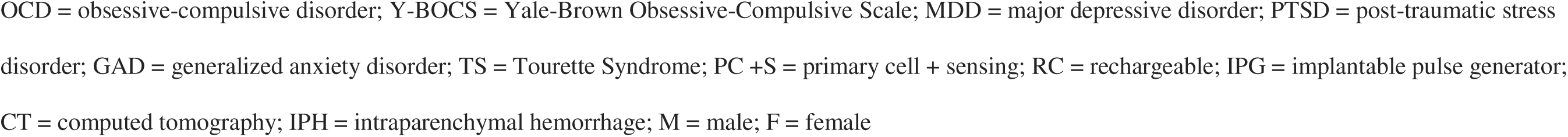
Demographics and past psychiatric history.

| *Patient* | *Gender* | *Age range* | *Race* | *Ethnicity* | *Comorbid diagnoses* | *OCD subtypes* | *Age range at OCD onset* | *Presurgical OCD Inpatient Treatment* | *Initial Y-BOCS-II (Y-BOCS)* | *Final Y-BOCS-II (Y-BOCS)* | *Adverse Events* |
| --- | --- | --- | --- | --- | --- | --- | --- | --- | --- | --- | --- |
| *P001* | M | 31-35 | White | Not Hispanic/Latino | MDD | Taboo thoughts, “just right” driven compulsions | 16-20 | Yes | 37 (35) | 8 (6) | PC+S reached end of life at Month 12, necessitating replacement with Activa RC and lack of stimulation for 48 hours |
| *P002* | F | 36-40 | White | Hispanic/Latino | PTSD, MDD, GAD | Contamination, Harm to Others, Neutralizing Compulsions | 16-20 | No | 39 (34) | 11 (9) | Mid-study IPG repositioning due to discomfort |
| *P003* | F | 36-40 | White | Hispanic/Latino | PTSD, MDD, TS | Harm to loved ones, repeating “just right” compulsions | 11-15 | No | 49 (39) | 28 (26) |  |
| *P004* | M | 36-40 | White | Not Hispanic/Latino | MDD, Bipolar Disorder II | Contamination, dirt and germs, washing, avoidance | 16-20 | No | 42 (38) | 25 (22) |  |
| *P005* | F | 31-35 | White | Not Hispanic/Latino | MDD | Contamination, harm to others, checking | 6-10 | Yes | 47 (37) | 14 (12) |  |
| *P006* | M | 36-40 | Asian | Not Hispanic/Latino | GAD | Intrusive unwanted thoughts, repeating, “just right” compulsions | 30-35 | No | 37 (35) | 0 (0) |  |
| *P007* | M | 36-40 | White | Not Hispanic/Latino | MDD | Harm to others, magical thinking, repeating until “just right” | 11-15 | No | 45 (36) | 21 (20) | Mid-study IPG repositioning due to discomfort and migration. Headache requiring admission; self-resolved. |
| *P008* | F | 36-40 | White | Not Hispanic/Latino | MDD, GAD | Contamination, harm to others, mental rituals and checking | 6-10 | Yes | 37 (35) | 1 (1) |  |
| *P009* | F | 26-30 | White | Hispanic/Latino | MDD, GAD | Taboo thoughts, scrupulosity, harm to others, multiple rituals | 11-15 | No | 45 (40) | 26 (24) |  |
| *P010* | M | 31-35 | White | Not Hispanic/Latino | MDD | Disgust contamination, extensive washing, marked avoidance | 11-15 | Yes | 46 (38) | 32 (31) | Postoperative CT revealed small IPH at the inferior left frontal lobe, patient remained asymptomatic and the IPH was self-limited |

OCD = obsessive-compulsive disorder; Y-BOCS = Yale-Brown Obsessive-Compulsive Scale; MDD = major depressive disorder; PTSD = post-traumatic stress disorder; GAD = generalized anxiety disorder; TS = Tourette Syndrome; PC +S = primary cell + sensing; RC = rechargeable; IPG = implantable pulse generator; CT = computed tomography; IPH = intraparenchymal hemorrhage; M = male; F = female

### Electrode Placement and Stimulation Parameters

All patients received a pair of DBS leads targeting the VC. Five patients were implanted in the bilateral VC/BNST, 4 in the bilateral VC/VS, and 1 in a mixed configuration (Figure 1a; Suppl. Table 1). In the final 5 subjects (P006-010), we also implanted a pair of electrocorticography (ECoG) strip electrodes over the OFC for recording only. The first 2 patients received a single Medtronic PC+S IPG, the next 3 (P003-005) received a single RC+S, and the last 5 (P006-010), who had the additional pair of ECoG strips, received 2 RC+S IPGs.

**Figure 1.**
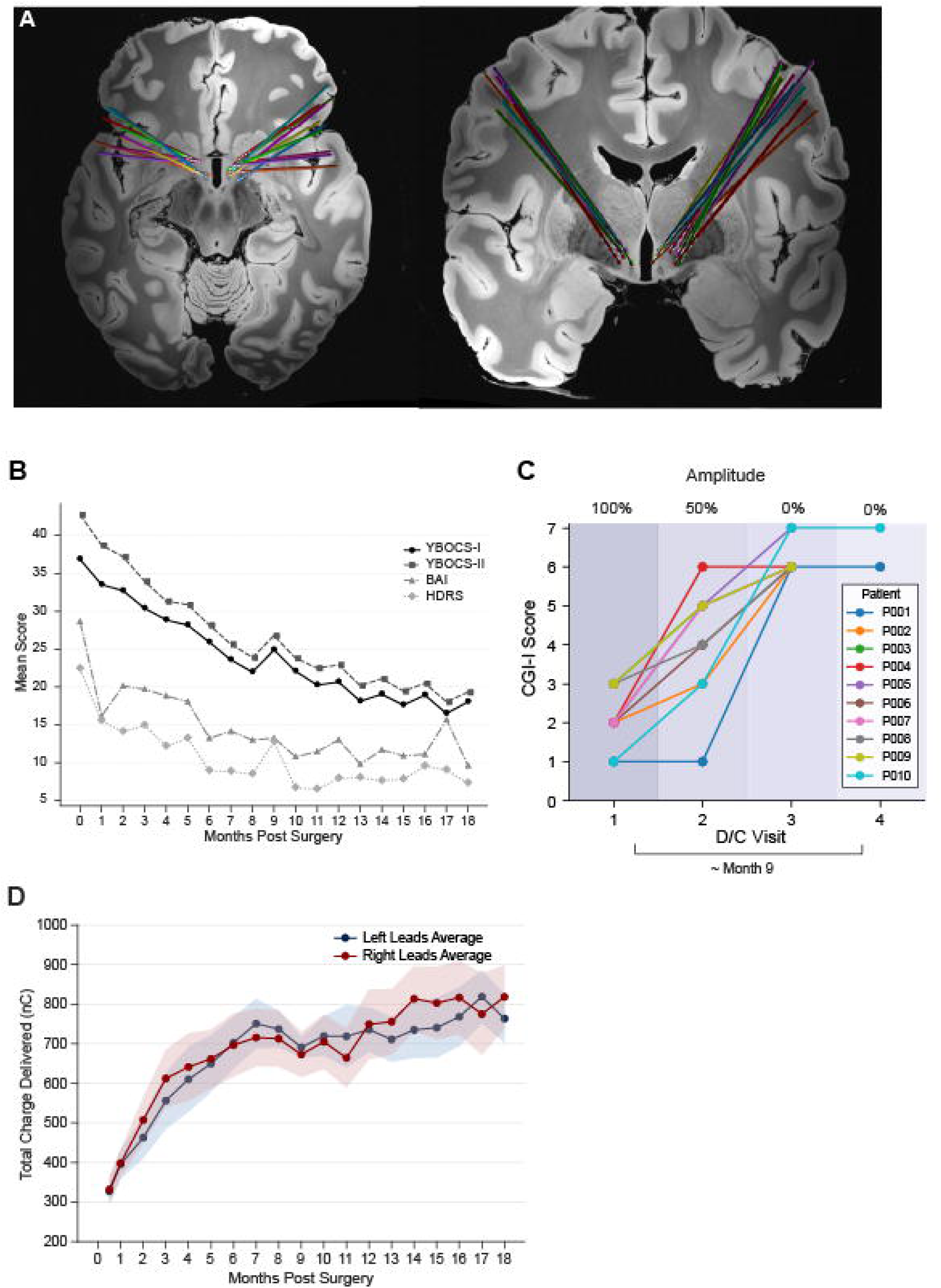
DBS lead locations, clinical scores over the course of the study, and total charge delivered through the DBS leads. **A)** Axial and coronal T1-weighted MRIs showing all 10 participants’ DBS leads terminating in the region of the ventral capsule (VC), either anterior to the anterior commissure (AC) such that the lead tip borders the ventral striatum (VC/VS), or posterior to the AC, such that the lead tip borders the bed nucleus of the stria terminalis (VC/BNST). **B)** Mean clinical scores (Y-BOCS, Y-BOCS-II, BAI, and HDRS) for all patients decreased during open-label stimulation (months 1-8) and then again in open-label continuation (months 10-18). **C)** During double-blind discontinuation at month 9, stimulation amplitude was tapered according to a fixed schedule: 100% during week 1, 50% during week 2, and 0% stimulation for weeks 3 and 4. All patents experienced increased CGI-I scores, indicating true rather than sham response over the preceding months. **D)** The mean total charge delivered increased across the left (blue) and right (red) DBS leads over the course of DBS programming. Shaded regions indicate the standard error of the mean.

All patients received monopolar stimulation, most on one of the middle contacts. Final stimulation parameters and MNI coordinates of the stimulating contact are shown in Supplemental Table 1. For those leads (16/20) that were in a sensing-compatible configuration, we recorded local field potentials on flanking contacts with bipolar referencing.

### OCD Outcomes

The trial lasted 18 months and had 4 phases. Phase 1 (months 0-6) consisted of open-label programming to identify optimal stimulation parameters. During months 7-8 (Phase 2), patients underwent ERP(23) to test whether ERP added to ongoing DBS could provide further clinical benefit. We then performed double-blind discontinuation of DBS therapy (month 9; Phase 3) to distinguish true response from sham response. Finally, patients entered an open-label follow-up period (months 10-18; phase 4) for the remainder of the study.

Consistent with previous convention(24), we defined clinical response as a ≥35% decrease in Y-BOCS and a partial response as a 25-34% decrease. At the end of phase 1 (6 months), the mean reduction in Y-BOCS was 13 points or 36% (Table 2), or 17 points or 41% on Y-BOCS-II (Supp. Table 2). Clinical response rate at this time point was 60% (6/10), with no additional partial responders.

**Table 2.** Y-BOCS and percent reduction in scores across the study, and Changes in Clinical Global Impression-Improvement (CGI-I) across the withdrawal phase.

| Patient | Pre-Surgical<br>Baseline | Y-BOCS |  |  |  |  |  | CGI-I |  |  |  |
| --- | --- | --- | --- | --- | --- | --- | --- | --- | --- | --- | --- |
| | | 6 Months | | 9 Months | | 18 Months | | Pre | Post | $\Delta$ | D/C Phase<br>Duration (d) |
|  |  | Score | %<br>Reduction | Score | %<br>Reduction | Score | %<br>Reduction |  |  |  |  |
| P001 | 35 | 19 | 45.7 | 18 | 48.6 | 6 | 82.9 | 1 | 6 | 5 | 13 |
| P002 | 34 | 15 | 55.9 | 15 | 55.9 | 9 | 73.5 | 2 | 6 | 4 | 11 |
| P003 | 39 | 36 | 7.7 | 37 | 5.1 | 26 | 33.3 | 3 | 6 | 3 | 9 |
| P004 | 38 | 24 | 36.8 | 29 | 23.7 | 22 | 42.1 | 2 | 6 | 4 | 11 |
| P005 | 37 | 30 | 18.9 | 26 | 29.7 | 12 | 67.6 | 2 | 6 | 4 | 7 |
| P006 | 35 | 18 | 48.6 | 9 | 74.3 | 0 | 100.0 | 1 | 6 | 5 | 10 |
| P007 | 36 | 22 | 38.9 | 27 | 25.0 | 20 | 44.4 | 2 | 6 | 4 | 6 |
| P008 | 35 | 7 | 80.0 | 7 | 80.0 | 1 | 97.1 | 1 | 6 | 5 | 13 |
| P009 | 40 | 33 | 17.5 | 32 | 20.0 | 24 | 40.0 | 3 | 6 | 3 | 23 |
| P010 | 38 | 33 | 13.2 | 33 | 13.2 | 31 | 18.4 | 4 | 7 | 3 | 18 |
| Mean (± SD) | 36.7 (2.0) | 23.7 (9.3) | 36.3 (22.5) | 23.3 (10.4) | 37.4 (25.8) | 15.1 (10.9) | 59.9 (28.2) | | | | |

CGI-I = Clinical Global Impression- Improvement; D/C = discontinuation; d = number of days; $\Delta$ = change in score

The phase 2 ERP booster consisted of 15 sessions of twice-weekly outpatient ERP delivered over 2 months. Mean Y-BOCS change during this phase was a 3.7 point decrease (3.9 point decrease in Y-BOCS-II; Table 3). At an individual patient level, 6 patients decreased by more than 1 point on Y-BOCS, compared to only one who increased by more than 1 point.

**Table 3.** Comparison of baseline, pre-ERP, and post-ERP Y-BOCS-II.

| Patient | Y-BOCS |  |  |  |  |  | YBOCS-II |  |  |  |  |  |
| --- | --- | --- | --- | --- | --- | --- | --- | --- | --- | --- | --- | --- |
| | Pre-Surgical Baseline | 2 months pre-ERP | Immediately pre-ERP | Post-ERP | ΔPre-ERP<sup>1</sup> | ΔERP<sup>2</sup> | Pre-Surgical Baseline | 2 months pre-ERP | Immediately pre-ERP | Post-ERP | ΔPre-ERP<sup>1</sup> | ΔERP<sup>2</sup> |
| P001 | 35 | 18 | 19 | 15 | +1 | -4 | 37 | 20 | 21 | 17 | +1 | -4 |
| P002 | 34 | 14 | 15 | 13 | +1 | -2 | 39 | 15 | 17 | 15 | +2 | -2 |
| P003 | 39 | 37 | 36 | 37 | -1 | +1 | 49 | 38 | 39 | 40 | +1 | +1 |
| P004 | 38 | 24 | 23 | 22 | -1 | -1 | 42 | 26 | 25 | 23 | +1 | -2 |
| P005 | 37 | 31 | 30 | 22 | -1 | -8 | 47 | 34 | 34 | 25 | 0 | -9 |
| P006 | 35 | 19 | 18 | 3 | -1 | -15 | 37 | 20 | 19 | 4 | -1 | -15 |
| P007 | 36 | 35 | 22 | 25 | -13 | +3 | 45 | 39 | 25 | 26 | -14 | +1 |
| P008 | 35 | 6 | 6 | 4 | 0 | -2 | 37 | 7 | 8 | 6 | +1 | -2 |
| P009 | 40 | 33 | 33 | 24 | 0 | -9 | 45 | 34 | 34 | 27 | 0 | -7 |
| P010 | 38 | 31 | 33 | 33 | +2 | 0 | 46 | 33 | 34 | 34 | +1 | 0 |
| Mean (± SD) | 36.7 (2.0) | 24.8 (10.3) | 23.5 (9.5) | 19.8 (11.2) | -1.3 (4.2) | -3.7 (5.5) | 42.4 (4.6) | 26.6 (10.8) | 25.6 (9.7) | 21.7 (11.4) | -0.8 (4.7) | -3.9 (5.1) |

Note. <sup>1</sup>Change in scores from 2 months prior to the ERP intervention period to the final pre-ERP assessment. <sup>2</sup>Change in scores from the final pre-ERP assessment to the first post-ERP assessment, spanning approximately 2 months and 15 ERP sessions; ERP = exposure-response prevention therapy; Y-BOCS (II) = Yale-Brown Obsessive-Compulsive Scale (Second Edition); SD = standard deviation

**Table 4.** HDRS outcomes prior to discontinuation (D/C) phase and at end of study relative to pre-surgical baseline.

| *Patient* | *Prior to D/C Phase (Month 9)* |  |  |  | *End of Study (Month 18)* |  |  |
| --- | --- | --- | --- | --- | --- | --- | --- |
|  | **Pre-Surgical Baseline** | **Score** | **Δ HDRS** | **% Reduction** | **Score** | **Δ HDRS** | **% Reduction** |
| *P001* | 12 | 3 | -9 | 75.0 | 2 | -10 | 83.3 |
| *P002* | 26 | 17 | -9 | 34.6 | 10 | -16 | 61.5 |
| *P003* | 31 | 16 | -15 | 48.4 | 3 | -28 | 90.3 |
| *P004* | 16 | 10 | -6 | 37.5 | 10 | -6 | 37.5 |
| *P005* | 27 | 3 | -24 | 88.9 | 3 | -24 | 88.9 |
| *P006* | 19 | 1 | -18 | 94.7 | 1 | -18 | 94.7 |
| *P007* | 28 | 15 | -13 | 46.4 | 4 | -24 | 85.7 |
| *P008* | 18 | 1 | -17 | 94.4 | 1 | -17 | 94.4 |
| *P009* | 20 | 4 | -16 | 80.0 | 8 | -12 | 60.0 |
| *P010* | 26 | 18 | -8 | 30.8 | 15 | -11 | 42.3 |
| *Mean (± SD)* | 22.3 (6.1) | 8.8 (7.1) | -13.5 (5.6) | 63.1 (26.0) | 5.7 (4.8) | -16.6 (7.1) | 73.9 (21.7) |

Notable examples are P001, P005, P009, and P006, whose Y-BOCS decreased by 4, 8, 9, and 15 points, respectively (Table 3). Respective numbers on Y-BOCS-II were 7 patients who decreased by more than 1 point, and 0 who increased by more than 1 point. It is possible that severity scores would decrease over these 2 months simply because of ongoing stimulation and not because of the ERP addition. To account for this possibility, we compared the change during the 2-month ERP phase with the change during the 2 months prior to the ERP phase (i.e., months 5-6). This comparison is not perfect because the effect of stimulation may vary month to month, but it provides a rough estimate of the contribution of ERP. Mean change in the 2 prior months was a decrease of 1.3 points on Y-BOCS (0.8 on Y-BOCS-II), with only 1 patient decreasing by more than 1 point and 1 increasing by more than 1 point on Y-BOCS (1 and 1 respectively on Y-BOCS-II as well). Comparing the 2 time points within individuals, 7 patients fared better during the ERP period than during the previous 2-month period (the 4 mentioned above and P002, P008, P010; Table 3), 2 fared worse, and 1 was unchanged based on Y-BOCS. These individual-level effects were not large or consistent enough to produce a significant group effect using Y-BOCS scores (p=0.18, binomial test between pairs). Using Y-BOCS-II, however, 8 patients improved more during the ERP period than during the preceding 2-month period, 1 worsened more, and 1 was unchanged. A binomial test of these group results did show a significant difference (p=0.04). As there is no a priori reason to prefer one scale over the other for this assessment, the group results suggest that ERP may have provided additional benefit over DBS, but not with a consistent effect. Individual-level effects, on the other hand, were present.

By the 9-month time point, just prior to the Phase 3 blinded discontinuation, the mean reduction in Y-BOCS across the cohort was 13 points or 37% (Table 2). Four of the 10 patients (40%) were responders, and 2 more were partial responders. By Y-BOCS-II, the mean reduction was 17 points or 43%, 6 patients (60%) were responders, and 1 was a partial responder (Supp. Table 2).

During the discontinuation, clinical visits were increased in frequency to weekly. Patients were told that their stimulation would be reduced but were not told when the change would occur to reduce a nocebo effect. Raters were also blinded to stimulation settings. None of the patients tolerated withdrawal of DBS therapy. Table 2 and Figure 1C indicate CGI-I scores during this period and demonstrate the increase across all patients, as well as the duration spent in this phase before rescue with resumption of stimulation. Non-tolerance of withdrawal in those who had responded by this time indicates true (rather than sham) response. Interestingly, even those who were non-responders at this time (e.g., P003, P010) did not tolerate reduction/cessation of therapy. Symptoms produced during withdrawal were anxiety, depressed mood, sense of doom, etc., not OCD symptoms per se.

All 10 patients elected to resume stimulation after the discontinuation and continued into Phase 4, open-label follow-up. Clinical improvement continued (Fig. 1B), and by the end of the study, mean Y-BOCS reduction was 22 points or 60%, with 8 full responders, 1 partial responder, and 1 non-responder (Table 2). On Y-BOCS-II, scores decreased by 26 points or 63%, with 9 full responders and 1 partial responder (Supp. Table 2).

### Other Psychiatric and Neuropsychological Outcomes

Depression and anxiety scores also decreased over the course of the trial. As shown in Figure 1B, these metrics decreased more rapidly than did OCD scores following DBS activation. Prior to phase 3 (month 9), the Hamilton Depression Rating Scale (HDRS) decreased by 14 points or 63%. Using the standard criterion for full response as 50% reduction, 5 patients (50%) had achieved full response in co-morbid depression by this time point. By the end of study (month 18), HDRS had decreased by 17 points or 74%, and 8 patients (80%) were full responders. Using a threshold of HDRS<7 to define remission, 5 patients (50%) had achieved remission of their depressive symptoms by month 9, and 6 (60%) by month 18.

The occurrence of (hypo)mania was determined by a combination of clinical judgement of W.K.G and scores on the Young Mania Rating Scale (YMRS), using 13-19 for hypomania and ≥20 for mania. By these criteria, one subject developed mania (YMRS=28) and one subject became hypomanic (YMRS=17) during DBS treatment. Both responded to temporary lowering of stimulation amplitude and co-administration of mood stabilizers. They both became and stayed euthymic after these interventions.

Across all cognitive domains examined, analyses did not detect statistically significant differences between baseline and long-term post-surgical assessments (all p-values>0.05).

Results revealed a statistically significant reduction in symptoms of depression as measured by the Beck Depression Inventory-Second Edition (BDI-II) from baseline to long-term post-surgical follow-up (p=0.020). Mean BDI-II scores decreased from 38.2 at baseline to 19.2 at long-term post-surgical assessment. We observed a statistically significant reduction in worry as measured by the Penn State Worry Questionnaire from baseline to long-term follow-up (p=0.034); total scores decreased from a mean of 74.8 to 57.7. Additionally, significant reductions were observed across multiple OCD symptom dimensions on the OCI-R. Washing symptoms decreased from a mean of 7.00 at baseline to 3.56 at long-term post-surgical follow-up (p=0.008). Checking symptoms likewise declined (6.56 to 1.89; p=0.005), as did obsessing symptoms (10.67 to 5.44; p=0.005).

### Adverse Events

One patient had a small (∼2 cc) intraparenchymal hematoma in the left OFC, presumably caused by the ECoG strip during the placement procedure (Table 1). The hematoma was clinically silent and self-limited. Two patients underwent an outpatient procedure to reposition an IPG due to discomfort. One patient experienced unexpected battery depletion in their PC+S IPG resulting in loss of therapy for 48 hours until the IPG was replaced. There were no infections or clinically significant hemorrhages.

## Discussion

We report 3 main clinical findings. First, 9 of the 10 patients achieved at least partial response criterion (8 full response, 1 partial) in the open-label phase based on Y-BOCS. Applying the same response thresholds for the Y-BOCS-II, which provides more granular differentiation at the severe end of the scale, we had 9 full responders and 1 partial responder. All responders had to be rescued during blinded discontinuation, indicating that the responses were likely true, not sham. Second, ERP was tolerated by all patients and provided additional benefit to several individuals despite its ineffectiveness prior to DBS. Third, secondary outcomes of depression, anxiety, and disability improved significantly.

Our clinical outcomes of 80% response rate (90% if using Y-BOCS-II) compare favorably to those of previous trials targeting the VC region. Goodman et al.(10) showed a 67% response rate, Denys et al.(12) 56%, and Greenberg et al.(11) 62%. A Belgian group, who published the first study of DBS for OCD in 1999(25), reported in 2015 on 24 patients using an open-label optimization followed by blinded crossover design, with a 67% response rate at long-term (median 6 years) follow-up. Most recently, Mosley et al.(15) used an up-front randomization to blinded active vs. sham design followed by open-label stimulation in 9 patients. During the blinded phase, the active group improved significantly relative to sham, and in the open-label continuation, 78% responded.

Aggregating over the published literature (including other DBS targets), the meta-analytic response rate was 66% across 352 patients in 34 studies from 2005 to 2021(17). The large number of well-controlled trials demonstrating significant improvement with VC DBS supports the consensus guidelines recommendation to consider this therapy for otherwise treatment-refractory OCD patients(26).

Our participants engaged in an ERP booster course approximately 6 months after initiating DBS. Despite unresponsiveness to prior ERP as a criterion for trial inclusion, several patients gained additional benefit from ERP following DBS. Seven of our 10 trial participants obtained a further reduction in Y-BOCS during the 2-month ERP booster (8 by Y-BOCS-II criteria). Other trials have observed a similar synergistic response between DBS and ERP(12,20,27,28). Our previous neurophysiological studies in a non-overlapping cohort of DBS for OCD patients shed some light on a possible mechanism explaining this DBS-ERP synergy(18,20,27). During ERP, patients gradually and systematically confront distressing stimuli while abstaining from compulsions and/or avoidant behaviors. Over time, as the feared consequences underlying compulsions and/or avoidance do not manifest (i.e., the calamitous event does not occur if a phrase is not repeated a certain number of times), individuals learn a new, non-threatening relationship with previously triggering stimuli. Rather than avoiding these external or internal stimuli, individuals thus gradually learn to approach and tolerate them(29). Our working model(18) conceives of VC DBS as a strong driver of this pro-approach diathesis, facilitating transition from an avoidant to an approachful phenotype. The subcortical and prefrontal cortical regions engaged by VC stimulation (nucleus accumbens, OFC, ventromedial and ventrolateral prefrontal cortex) are known to regulate the reward-motivated behavior whose modulation may underlie this transition. This putative common mechanism behind ERP and DBS may explain their synergistic interaction.

Indeed, overstimulation of this target can drive approach behavior into a disinhibited regime characterized by recklessness, overspending, decreased need for sleep, and increased libido. This stimulation-related side effect has been commonly observed in prior trials(28,30,31) and fits within the predictions of our approach-avoidance model(18) as an effect of unbridled approach behavior. This observation has practical consequences for DBS programming and patient management. Our general strategy for programming this target is to increase stimulation over successive clinic visits while being mindful not to enter this overstimulation regime, which acutely may manifest as increased talkativeness, restlessness, or energy. Over time, this stimulation ceiling increases, such that settings that were not tolerated earlier in therapy are not only tolerated but therapeutic later. The steady increase in charge delivery (by increasing either amplitude or pulse width) we measured in this cohort (Figure 1D) illustrates this pattern.

This approach-avoidance framework offers additional benefits in terms of informing patient selection and surgical targeting that may have contributed to our favorable outcomes. The OCD subtypes characterized by harm avoidance may respond optimally to VC DBS, given the tendency of stimulation here to drive approach behavior. Examples include unwanted aggressive thoughts accompanied by rituals driven by a fear that failing to perform the compulsions would result in harm to loved ones. The contamination subtype driven by a concern that contaminant exposure would cause harm to self or others is another example. In contrast, contamination OCD associated with the feeling of “disgust” in reaction to the offending substance but without a fear of harmful consequences may suggest a different underlying mechanism(32) that may not respond as readily to VC stimulation. Indeed, this disgust phenotype is also notoriously difficult to treat with ERP(33). Consistent with this framework and with historical ERP experience, the single clear non-responder in our cohort (P010) had a disgust contamination subtype, whereas all others demonstrated harm avoidance and improved. DBS targeting the subthalamic nucleus or globus pallidus internus for PD is only offered to patients with motor symptoms that are known to respond to stimulation of these targets (i.e., tremor, bradykinesia, rigidity) and not to those with predominantly non-responsive motor symptoms (i.e., gait instability, freezing). In the same way, DBS for OCD will greatly benefit from understanding which symptoms improve and selecting candidates accordingly.

This framework also impacts our surgical targeting strategy. As we have previously described(13) we use intraoperative awake testing to identify approach behavior and place the lead in the location that produces this effect. This combination of patient selection and targeting, along with the programming strategy mentioned above, may optimally match circuit pathology (i.e., approach-avoidance framework) with therapy delivery and may have contributed to our promising response rate.

The transition from short-term effects (increased approach behavior) to improvements in OCD symptoms per se likely reflects plasticity in the cortico-basal circuits governing reward-motivated decision-making behavior and may be associated with measurable neurophysiological effects. We have previously used the recording capacity of this investigational DBS platform (Medtronic Activa PC+S and Summit RC+S) to study neural correlates of reward-motivated behavior in a subset of these trial patients(16,34). Others have used this system for similar investigations in PD(19,35), chronic pain(36), and depression(37). Although these investigational devices are no longer being manufactured, a commercially available DBS platform with recording capability (Medtronic Percept) has enabled continued neurobehavioral research. Its design for band-power estimation is proving useful for closed-loop/adaptive applications in PD(38), but the time course of symptom evolution and neural mechanism in OCD may require a different approach(18,39).

All 10 participants enjoyed reduced depression severity, with a mean HDRS reduction of 16.6 points (74%) across the cohort by the end of the study. Eight of the 10 achieved full response criterion (≥50% reduction), and 6/10 achieved remission (HDRS<7). Even our single non-responder, whose Y-BOCS only decreased by 18%, experienced a 42% improvement in depression severity. This improvement in co-morbid depression has been documented in previous DBS trials for OCD(12,14,15). Much less common, however, is analysis of depression outcome stratified by its severity at enrollment. An HDRS threshold of ≥20 is often used as the severity criterion for trials of treatment-resistant depression (TRD). Six of our participants met that criterion at enrollment, indicating high-moderate to severe co-morbid depression. Of these 6, 5 achieved full response criterion, and 3 were remitters. We do not suggest that these 6 were identical to individuals with primary TRD, as there are important differences in terms of diagnostic criteria, previous therapies, etc. But we highlight the point that DBS of this region may be worth considering even for treating a significant co-morbidity, beyond its primary indication alone.

## Disclosures

W.K.G. receives research support from Abbott Laboratories and royalties from Nview, LLC and OCDscales, LLC. S.A.S. is a consultant for Boston Scientific, Abbott, Neuropace, Koh Young, and Zimmer Biomet, and co-founder of Motif Neurotech. E.A.S. receives direct funding from the International OCD Foundation as well as MHNTI for providing trainings on treating obsessive-compulsive disorder with psychotherapy. He was a consultant for Brainsway and Biohaven Pharmaceuticals in the past 36 months. He owns stock options less than $5000 in NView (for distribution of the Y-BOCS and CY-BOCS) and receives royalties from OCD Scales LLC (for distribution of the Y-BOCS and CY-BOCS). He receives book royalties from Elsevier, Wiley, Oxford, American Psychological Association, Guildford, Springer, Routledge, and Jessica Kingsley.

## Data Availability

All data produced in the present study are available upon reasonable request to the authors.

## Acknowledgements

The authors thank the participants and their families for their involvement in the research program. Activa PC+S and Summit RC+S devices were donated by Medtronic as part of the BRAIN Initiative Public-Private Partnership Program. The research was supported by the National Institutes of Health (NIH) NINDS BRAIN Initiative via contracts UH3NS100549 (to W.K.G, S.A.S. E.A.S., N.R.P.) and by the McNair Foundation (S.A.S., N.R.P.), Gordon and Mary Cain Pediatric Neurology Research Foundation (S.A.S.), Ream Foundation (E.A.S.), and International OCD Foundation (E.A.S.).

## Notes

### Clinical Trial

NCT04281134

### Funding Statement

The research was supported by the National Institutes of Health NINDS BRAIN Initiative via contracts UH3NS100549, by the McNair Foundation, and by the Gordon and Mary Cain Pediatric Neurology Research Foundation. Medtronic donated the DBS devices and related hardware.

### Author Declarations

The institutional review board of Baylor College of Medicine gave ethical approval for this work.

## References

1. Rasmussen SA, Eisen JL. The Epidemiology and Clinical Features of Obsessive Compulsive Disorder. Psychiatr Clin North Am. 1992;15(4):743–758. doi:10.1016/s0193-953x(18)30205-3

2. Ruscio AM, Stein DJ, Chiu WT, Kessler RC. The epidemiology of obsessive-compulsive disorder in the National Comorbidity Survey Replication. Mol Psychiatry. 2010;15(1):53–63. doi:10.1038/mp.2008.94

3. Meier SM, Mattheisen M, Mors O, Schendel DE, Mortensen PB, Plessen KJ. Mortality Among Persons With Obsessive-Compulsive Disorder in Denmark. JAMA Psychiatry. 2016;73(3):268. doi:10.1001/jamapsychiatry.2015.3105

4. Deacon BJ, Abramowitz JS. Cognitive and behavioral treatments for anxiety disorders: A review of meta analytic findings. J Clin Psychol. 2004;60(4):429–441. doi:10.1002/jclp.10255

5. Goodman WK, Price LH, Delgado PL, et al. Specificity of Serotonin Reuptake Inhibitors in the Treatment of Obsessive-Compulsive Disorder: Comparison of Fluvoxamine and Desipramine. Arch Gen Psychiatry. 1990;47(6):577–585. doi:10.1001/archpsyc.1990.01810180077011

6. Öst LG, Havnen A, Hansen B, Kvale G. Cognitive behavioral treatments of obsessive–compulsive disorder. A systematic review and meta-analysis of studies published 1993–2014. Clin Psychol Rev. 2015;40:156–169. doi:10.1016/j.cpr.2015.06.003

7. Simpson HB, Huppert JD, Petkova E, Foa EB, Liebowitz MR. Response Versus Remission in Obsessive-Compulsive Disorder. J Clin Psychiatry. 2006;67(02):269–276. doi:10.4088/jcp.v67n0214

8. Eisen JL, Goodman WK, Keller MB, et al. Patterns of Remission and Relapse in Obsessive-Compulsive Disorder: A 2-Year Prospective Study. J Clin Psychiatry. 1999;60(5):346–351. doi:10.4088/jcp.v60n0514

9. Dunlop K, Woodside B, Olmsted M, Colton P, Giacobbe P, Downar J. Reductions in Cortico-Striatal Hyperconnectivity Accompany Successful Treatment of Obsessive-Compulsive Disorder with Dorsomedial Prefrontal rTMS. Neuropsychopharmacology. 2016;41(5):1395–1403. doi:10.1038/npp.2015.292

10. Goodman WK, Foote KD, Greenberg BD, et al. Deep Brain Stimulation for Intractable Obsessive Compulsive Disorder: Pilot Study Using a Blinded, Staggered-Onset Design. Biol Psychiatry. 2010;67(6):535–542. doi:10.1016/j.biopsych.2009.11.028

11. Greenberg BD, Gabriels LA, Malone DA, et al. Deep brain stimulation of the ventral internal capsule/ventral striatum for obsessive-compulsive disorder: worldwide experience. Mol Psychiatry. 2010;15(1):64–79. doi:10.1038/mp.2008.55

12. Denys D, Mantione M, Figee M, et al. Deep Brain Stimulation of the Nucleus Accumbens for Treatment-Refractory Obsessive-Compulsive Disorder. Arch Gen Psychiatry. 2010;67(10):1061–1068. doi:10.1001/archgenpsychiatry.2010.122

13. Shofty B, Gadot R, Viswanathan A, et al. Intraoperative valence testing to adjudicate between ventral capsule/ventral striatum and bed nucleus of the stria terminalis target selection in deep brain stimulation for obsessive-compulsive disorder. J Neurosurg. Published online 2022:1–9. doi:10.3171/2022.10.jns221683

14. Luyten L, Hendrickx S, Raymaekers S, Gabriëls L, Nuttin B. Electrical stimulation in the bed nucleus of the stria terminalis alleviates severe obsessive-compulsive disorder. Mol Psychiatry. 2016;21(9):1272–1280. doi:10.1038/mp.2015.124

15. Mosley PE, Windels F, Morris J, et al. A randomised, double-blind, sham-controlled trial of deep brain stimulation of the bed nucleus of the stria terminalis for treatment-resistant obsessive-compulsive disorder. Transl Psychiatry. 2021;11(1):190. doi:10.1038/s41398-021-01307-9

16. Provenza NR, Sheth SA, Rijn EMD van, et al. Long-term ecological assessment of intracranial electrophysiology synchronized to behavioral markers in obsessive-compulsive disorder. Nat Med. 2021;27(12):2154–2164. doi:10.1038/s41591-021-01550-z

17. Gadot R, Najera R, Hirani S, et al. Efficacy of deep brain stimulation for treatment-resistant obsessive-compulsive disorder: systematic review and meta-analysis. *J Neurol*, Neurosurg Psychiatry. 2022;93(11):1166–1173. doi:10.1136/jnnp-2021-328738

18. Provenza NR, Reddy S, Allam AK, et al. Disruption of neural periodicity predicts clinical response after deep brain stimulation for obsessive-compulsive disorder. Nat Med. Published online 2024:1–11. doi:10.1038/s41591-024-03125-0

19. Oehrn CR, Cernera S, Hammer LH, et al. Chronic adaptive deep brain stimulation versus conventional stimulation in Parkinson’s disease: a blinded randomized feasibility trial. Nat Med. Published online 2024:1–12. doi:10.1038/s41591-024-03196-z

20. Mantione M, Nieman DH, Figee M, Denys D. Cognitive–behavioural therapy augments the effects of deep brain stimulation in obsessive–compulsive disorder. Psychol Med. 2014;44(16):3515–3522. doi:10.1017/s0033291714000956

21. Miguel EC, Lopes AC, McLaughlin NCR, et al. Evolution of gamma knife capsulotomy for intractable obsessive-compulsive disorder. Mol Psychiatry. 2019;24(2):218–240. doi:10.1038/s41380-018-0054-0

22. Goodman WK, Price LH, Rasmussen SA, et al. The Yale-Brown Obsessive Compulsive Scale: I. Development, Use, and Reliability. Arch Gen Psychiatry. 1989;46(11):1006–1011. doi:10.1001/archpsyc.1989.01810110048007

23. Abramowitz JS, Foa EB, Franklin ME. Exposure and Ritual Prevention for Obsessive-Compulsive Disorder: Effects of Intensive Versus Twice-Weekly Sessions. J Consult Clin Psychol. 2003;71(2):394–398. doi:10.1037/0022-006x.71.2.394

24. Mataix Cols D, Cruz LF de la, Nordsletten AE, Lenhard F, Isomura K, Simpson HB. Towards an international expert consensus for defining treatment response, remission, recovery and relapse in obsessive compulsive disorder. World Psychiatry. 2016;15(1):80–81. doi:10.1002/wps.20299

25. Nuttin B, Cosyns P, Demeulemeester H, Gybels J, Meyerson B. Electrical stimulation in anterior limbs of internal capsules in patients with obsessive-compulsive disorder. Lancet. 1999;354(9189):1526. doi:10.1016/s0140-6736(99)02376-4

26. Staudt MD, Pouratian N, Miller JP, et al. Congress of Neurological Surgeons Systematic Review and Evidence-Based Guidelines for Deep Brain Stimulations for Obsessive-Compulsive Disorder: Update of the 2014 Guidelines. Neurosurgery. 2021;88(4):nyaa596-. doi:10.1093/neuros/nyaa596

27. Graat I, Mocking R, Figee M, et al. Long-term Outcome of Deep Brain Stimulation of the Ventral Part of the Anterior Limb of the Internal Capsule in a Cohort of 50 Patients With Treatment-Refractory Obsessive-Compulsive Disorder. Biol Psychiatry. 2021;90(10):714–720. doi:10.1016/j.biopsych.2020.08.018

28. Denys D, Graat I, Mocking R, et al. Efficacy of Deep Brain Stimulation of the Ventral Anterior Limb of the Internal Capsule for Refractory Obsessive-Compulsive Disorder: A Clinical Cohort of 70 Patients. Am J Psychiatry. 2020;177(3):265–271. doi:10.1176/appi.ajp.2019.19060656

29. Craske MG, Treanor M, Conway CC, Zbozinek T, Vervliet B. Maximizing exposure therapy: An inhibitory learning approach. Behav Res Ther. 2014;58:10–23. doi:10.1016/j.brat.2014.04.006

30. Widge AS, Licon E, Zorowitz S, et al. Predictors of Hypomania During Ventral Capsule/Ventral Striatum Deep Brain Stimulation. J Neuropsychiatry Clin Neurosci. 2016;28(1):38–44. doi:10.1176/appi.neuropsych.15040089

31. Greenberg BD, Malone DA, Friehs GM, et al. Three-Year Outcomes in Deep Brain Stimulation for Highly Resistant Obsessive–Compulsive Disorder. Neuropsychopharmacology. 2006;31(11):2384–2393. doi:10.1038/sj.npp.1301165

32. Shapira NA, Liu Y, He AG, et al. Brain activation by disgust-inducing pictures in obsessive-compulsive disorder. Biol Psychiatry. 2003;54(7):751–756. doi:10.1016/s0006-3223(03)00003-9

33. Mitchell BJ, Coifman KG, Olatunji BO. Is disgust more resistant to extinction than fear? A meta-analytic review of laboratory paradigms. Behav Res Ther. 2024;174:104479. doi:10.1016/j.brat.2024.104479

34. Provenza NR, Rajesh SV, Reyes G, et al. High beta power in the ventrolateral prefrontal cortex indexes human approach behavior: a case study. J Neurosci. Published online 2025:e1321242025. doi:10.1523/jneurosci.1321-24.2025

35. Gilron R, Little S, Perrone R, et al. Long-term wireless streaming of neural recordings for circuit discovery and adaptive stimulation in individuals with Parkinson’s disease. Nat Biotechnol. 2021;39(9):1078–1085. doi:10.1038/s41587-021-00897-5

36. Shirvalkar P, Prosky J, Chin G, et al. First-in-human prediction of chronic pain state using intracranial neural biomarkers. Nat Neurosci. 2023;26(6):1090–1099. doi:10.1038/s41593-023-01338-z

37. Alagapan S, Choi KS, Heisig S, et al. Cingulate dynamics track depression recovery with deep brain stimulation. Nature. 2023;622(7981):130–138. doi:10.1038/s41586-023-06541-3

38. Stanslaski S, Summers RLS, Tonder L, et al. Sensing data and methodology from the Adaptive DBS Algorithm for Personalized Therapy in Parkinson’s Disease (ADAPT-PD) clinical trial. npj Park’s Dis. 2024;10(1):174. doi:10.1038/s41531-024-00772-5

39. Sheth SA, Rolston JD, Goodman WK, Provenza NR. Is Psychiatry Ready for Closed-Loop Invasive Neuromodulation? JAMA Psychiatry. 2025;82(12). doi:10.1001/jamapsychiatry.2025.2710

