## Supplemental Tables 1 and 2 for "Early Feasibility Study of Sensing-enabled Ventral Capsule Deep Brain Stimulation in 10 Participants with Intractable Obsessive-Compulsive Disorder"

**Supplemental Table 1. Implant information, stimulation parameters, and MNI coordinates of active contacts at the end of the study.**

|  |  |  |  | **Stimulating Contact** | | **Sensing Contact** | | **Final Rate (Hz)** | | **Final Amplitude*** | | **Final Pulse Width (μs)** | | **Left Hemisphere** | | | **Right Hemisphere** | | |
| --- | --- | --- | --- | --- | --- | --- | --- | --- | --- | --- | --- | --- | --- | --- | --- | --- | --- | --- | --- |
| **Study Participant** | **DBS Device** | **DBS Target Region** | **B OFC Strip** | **L** | **R** | **L** | **R** | **L** | **R** | **L** | **R** | **L** | **R** | **x** | **y** | **z** | **x** | **y** | **z** |
| P001 | PC+S | B VC/BNST | N | 1-/C+ | 10-/C+ | 0-2 | 9-11' | 150 | 150 | 5.0 | 5.0 | 120 | 120 | -6.06 | -1.05 | -3.69 | 10.22 | 1.69 | -2.74 |
| P002 | PC+S | B VC/BNST | N | 2-/C+ | OFF | 1-3' | 9-11' | 150 | OFF | 6.0 | OFF | 120 | OFF | -8.68 | 1.57 | 0.21 | OFF | OFF | OFF |
| P003 | RC+S | B VCVS | N | 1-/C+ | 9-/C+ | 0-2 | 8-10' | 150.6 | 150.6 | 6.0 | 5.7 | 210 | 210 | -12.52 | 4.36 | -3.24 | 10.77 | 5.47 | -3.9 |
| P004 | RC+S | L VCVS/ R VC/BNST | N | 1-/C+ | 9-/C+ | 0-2 | 8-10' | 150.6 | 150.6 | 5.2 | 5.0 | 120 | 120 | -12.61 | 6.42 | -5.74 | 8.1 | 2.27 | -5.7 |
| P005 | RC+S | B VC/BNST | N | 1-/C+ | 9-/C+ | 0-2 | 8-10' | 150.6 | 150.6 | 5.2 | 5.7 | 180 | 180 | -5.53 | -1.04 | -6.46 | 8.3 | 0.07 | -6.18 |
| P006 | RC+S x2 | B VCVS | Y | 0-/C+ | 9-/C+ | NA | 8-10' | 150.6 | 150.6 | 5.5 | 5.5 | 120 | 150 | -8.39 | 5.07 | -5.04 | 10.93 | 4.53 | -4.18 |
| P007 | RC+S x2 | B VCVS | Y | 1-/C+ | 8-/C+ | 0-2 | NA | 150.6 | 150.6 | 5.8 | 4.3 | 180 | 180 | -13.11 | 4.37 | -3.11 | 8.36 | 3.19 | -4.53 |
| P008 | RC+S x2 | B VC/BNST | Y | 1-/C+ | 9-/C+ | 0-2 | 8-10' | 150.6 | 150.6 | 5.7 | 5.7 | 120 | 90 | -13.76 | 2.89 | 0.68 | 12.59 | 3 | -1.31 |
| P009 | RC+S x2 | B VC/BNST | Y | 0-/C+ | 8-/C+ | NA | NA | 150.6 | 150.6 | 5.5 | 5.5 | 120 | 120 | -7.33 | -0.11 | -3.42 | 8.68 | 2.35 | -4.07 |
| P010 | RC+S x2 | B VCVS | Y | 1-/C+ | 9-/C+ | 0-2 | 8-10' | 150.6 | 150.6 | 5.5 | 5.5 | 150 | 150 | -12.02 | 6.38 | -6.78 | 11.91 | 7.19 | -6.41 |
| Mean (± SD) |  |  |  |  |  |  |  |  |  | 5.6 (0.3) | 5.4 (0.5) | 144 (34.1) | 146.7 (38.1) |  |  |  |  |  |  |

*Final amplitude for P001 and P002 in V; remainder of patients in mA. Mean constant current amplitude (mA; P003-P010); DBS = deep brain stimulation; PC+S = primary cell + sensing; RC + S = rechargeable + sensing; B = bilateral; VC/BNST = ventral capsule/bed nucleus of the stria terminalis; VCVS = ventral capsule/ventral striatum; OFC = orbitofrontal cortex; C+ = case positive; Hz = Hertz; mA = milliamperes; μs = microseconds; L = left; R = right; N = No; Y = Yes; NA = not applicable; SD = standard deviation. MNI= Montreal Neurological Institute.

**Supplemental Table 2. Y-BOCS-II scores across the study.**

|  |  | **6 Months** | | **9 Months** | | **18 Months** | |
| --- | --- | --- | --- | --- | --- | --- | --- |
| **Patient** | **Pre-Surgical Baseline** | **Score** | **% Reduction** | **Score** | **% Reduction** | **Score** | **% Reduction** |
| P001 | 37 | 21 | 43.2 | 20 | 45.9 | 8 | 78.4 |
| P002 | 39 | 17 | 56.4 | 16 | 59.0 | 11 | 71.8 |
| P003 | 49 | 39 | 20.4 | 40 | 18.4 | 28 | 42.9 |
| P004 | 42 | 26 | 38.1 | 32 | 23.8 | 25 | 40.5 |
| P005 | 47 | 34 | 27.7 | 29 | 38.3 | 14 | 70.2 |
| P006 | 37 | 19 | 48.6 | 10 | 73.0 | 0 | 100.0 |
| P007 | 45 | 25 | 44.4 | 26 | 42.2 | 21 | 53.3 |
| P008 | 37 | 7 | 81.1 | 9 | 75.7 | 1 | 97.3 |
| P009 | 45 | 34 | 24.4 | 34 | 24.4 | 26 | 42.2 |
| P010 | 46 | 34 | 26.1 | 34 | 26.1 | 32 | 30.4 |
| Mean (± SD) | 42.4 (4.6) | 25.6 (9.9) | 41.1 (18.3) | 25 (10.7) | 42.7 (20.7) | 16.6 (11.4) | 62.7 (24.5) |

Y-BOCS-II=Yale-Brown Obsessive Compulsive Score (Second Edition); Δ = change in score; SD = standard deviation
